# Activities contributing to quality of life of nursing home residents with dementia: a survey study

**DOI:** 10.1101/2025.07.04.25330890

**Authors:** Marlon M.P. Smeitink, Miriam L. Haaksma, Hanneke J.A. Smaling, Wilco P. Achterberg, Monique A.A. Caljouw

## Abstract

**Aims:** To explore which activities are offered to nursing home residents with dementia, what their expected contribution to quality of life is, and who chooses and offers these activities.

**Design:** A cross-sectional survey.

**Methods:** Healthcare professionals, informal caregivers, and volunteers of nursing home residents with dementia were invited to participate via the nationwide networks of the Dutch Alzheimer Association, the six Dutch academic networks in care for older people, Alzheimer cafes, and the personal (social media) networks of the researchers in September/October 2021. Participants received a survey via e-mail, which included items about which activities are offered to nursing home residents with dementia, whether participants expected the activities to contribute to residents’ quality of life, and by whom the activities were offered. Descriptive statistics were used to analyze the data.

**Results:** In total, 129 activities were mentioned. The top five most mentioned activities were live music and concerts (n=144), walking (n=141), listening to music (n=133), celebrating national holidays (n=129), and talking about memories (n=125). A median proportion of 90% (IQR 75-97) of all activities was expected to contribute to the residents’ quality of life. Activities were offered by healthcare professionals, informal caregivers, and volunteers.

**Conclusion:** Many different activities are being offered to nursing home residents with dementia, but there is a discrepancy between activities that were expected to contribute to quality of life and activities that were offered most frequently. Hence, residents may benefit from improved tailoring of activities based on their expected contribution to quality of life. Further research into the facilitators and barriers to offering tailored activities in collaboration with caregivers is necessary to unravel why and how tailoring of activities should and can be improved.

**What is already known:**

- Activities contribute to the quality of life of nursing home residents with dementia
- Besides intervention programs, information about which activities are currently offered in nursing homes, and by whom, is unknown

**What this paper adds:**

- More than 129 activities are offered in Dutch nursing homes by both healthcare professionals, informal caregivers, and volunteers
- 90% of all activities was expected to contribute to the residents’ QoL
- Healthcare professionals, informal caregivers, and volunteers differ in their expectation of which activities contribute to the quality of life of residents with dementia

## Introduction

Dementia is a progressive disease for which there is no cure. Therefore, the main goal of caring for people living with dementia is to maintain and improve their quality of life and well-being [1-3]. Nursing homes are focused on optimizing the quality of life of their residents with dementia [3]. They apply a person-centered care approach, which includes conducting activities, prioritizing residents’ well-being, and improving the quality of relationships between professionals and persons with dementia [4, 5].

Psychosocial interventions, such as activities, can be used to optimize the quality of life of residents [6, 7]. Activities are of great importance for the quality of life of people with dementia, as they strengthen their relationship with their loved ones [8] and enable them to thrive in nursing homes [9]. An activity is defined as meaningful when it is enjoyable, engaging, suited to the individual’s preferences, abilities and skills, related to personally relevant goals, related to an aspect of their identity, or giving a sense of belonging [6, 10-12]. People with dementia indicate it is important that activities are person-centered, i.e. tailored to their individual needs and situation [6, 13], since this makes activities more meaningful for them [12]. In addition to contributing to the quality of life of nursing home residents with dementia, activities can also decrease passivity, agitation, disruptive behavior, apathy, and depression [12]. Tailored activities have shown to be effective in increasing mood, engagement [14], agitation, emotional well-being, and sleep quality [15].

Although several studies have shown the importance of activities for nursing home residents with dementia [1, 3, 11], a comprehensive overview of which activities are currently being offered in nursing homes, and by whom, is lacking. Knowledge about which activities are considered meaningful is necessary to provide information on how to improve quality of life [7, 16, 17]. This study aims to provide an overview of which activities are offered to people with dementia in Dutch nursing homes and by whom. Additionally, we present the expected contribution of those activities to quality of life of residents according to healthcare professionals, volunteers and informal caregivers.

## Methods

### Study design

An online survey was used as a fast and accessible way to gather information. This cross-sectional study was conducted in the Netherlands between September and October 2021.

### Survey development and content

The survey contained nine topics: (1) informed consent, (2) demographics of the participant, (3) demographics of the relative with dementia, (4) relationship between participant and their relative with dementia, (5) activities, (6) preferences of the participant’s relative with dementia, (7) people involved in activities, (8) implementation, (9) transition from home to nursing home. The current study used data from topics 1 to 7. The survey consisted of a base set of questions and a set of specific questions for each of the three participant groups. Questions could be multiple choice or open-ended. Questions regarding the extent to which activities are tailored to the residents’ individual wishes, needs and skills were scored on a VAS-scale (1 very bad - 10 very good). All questions and response options utilized for this study are shown in Supplementary Table S1.

The 16 activity categories used in the fifth survey topic were based on a literature search on activities for persons with dementia [7, 11, 12]. The following categories were included: art activities, animal activities, activities including dolls, exercise, sensory stimulation, cognitive stimulation, music and entertainment, reminiscence, outside activities, family and social activities, household activities, religious activities, activities focused on personal care, activities focused on holidays and events, aromatherapy, and activities focused on daily rituals. Participants were asked which activities were offered in the nursing homes via multiple choice questions. After selecting a variety of activity categories, follow-up questions were shown about the specific activities within each category. For example, within the category ‘art activities’ we asked: ‘’Which specific art activities are offered in the nursing home?’’. For each selected art activity participants were subsequently asked ‘’Do you expect this activity contributes to the quality of life of residents?’’. Every question regarding types of activities included the option ‘’other’’, with an open text field to enter additional (new) responses.

The survey was pilot tested by two healthcare professionals and one informal caregiver to ensure the questions were comprehensible and to test the survey completion time (approximately 25 minutes).

### Sampling and recruitment

Participants were recruited nationwide through convenience sampling. This was done via the Dutch Alzheimer Association, SANO (the six Dutch academic collaborative networks in care for older people) [18, 19], Alzheimer cafes (nationwide) and the personal (social media) networks of the researchers to reach as many people as possible. After signing up, participants received an email with a link to one of the three (group-specific) online versions of the survey, avoiding multiple participation.

Three groups of participants were eligible for this survey. The first group consisted of healthcare professionals. They had to be working in a nursing home with residents with dementia for at least 12 months. The second group included volunteers with at least 12 months experience as a volunteer, who contributed to activities for residents with dementia, and had at least monthly contact with residents with dementia. The third group of participants comprised informal caregivers. Informal caregivers were included when they had at least weekly contact with their relative with dementia who lived in a nursing home. All participants had to have knowledge of the Dutch language and be at least 18 years old to participate.

### Data analysis

All participants who completed the question ‘’*Which activities are offered in the nursing home where you work/your relative lives?’’* were included in this study, because this is the main question of the current study. Activities that were mentioned at least five times in the open text fields of ‘’other’’ were added to the overview of activities. Walking was an answer option in the exercise category but was also often mentioned under ‘’other’’ of the outside activities category. When the data were processed it was therefore decided to list walking as a separate category. Descriptive statistics were used to analyze the data using SPSS version 25 [20].

### Ethical considerations

The [blinded for review] Medical Ethics Committee declared that the Medical Research Involving Human Subjects Act did not apply to this study and provided a waiver of consent (protocol nr. N21.128). The anonymous survey data was stored in a secured environment which could only be accessed by authorized researchers.

## 3. Results

The sample of this study consisted of 207 participants; 91 healthcare professionals, 15 volunteers, and 101 informal caregivers, after exclusion of 15 participants due to missing data on questions regarding activities (Figure 1). The participants were mostly female (75.8%, n=157), born in the Netherlands (96.6%, n=200), and they were from all areas of the Netherlands, although most of them lived in the province of South-Holland. Demographics per participant group (healthcare professionals, volunteers and informal caregivers) are shown in Table 1.

**Table 1.**
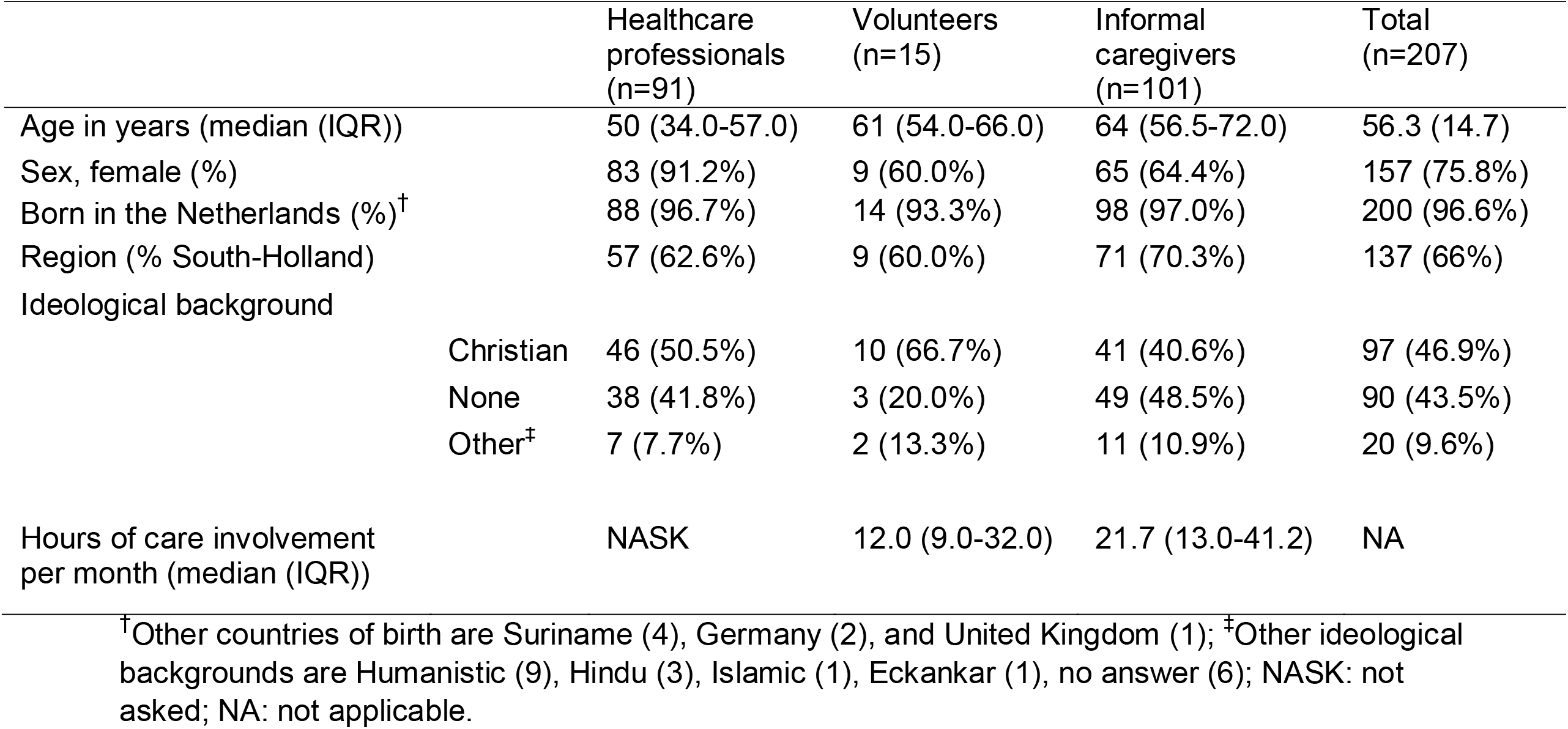
Demographics of the participants.

**Figure 1.**
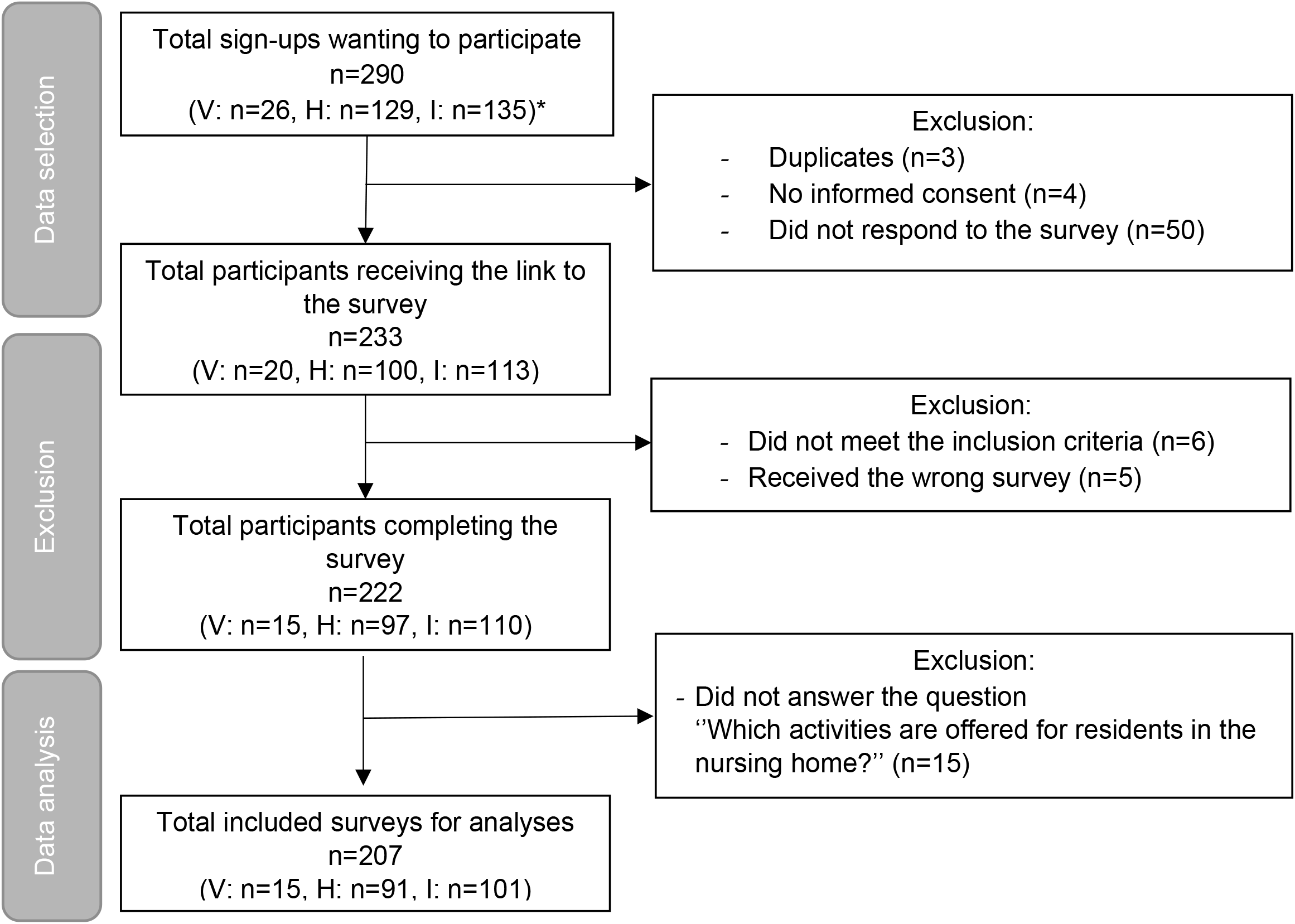
Flowchart of the surveys Note: *‘V’ Volunteers, ‘H’ Healthcare professionals, ‘I’ Informal caregivers.

Informal caregivers had a median age of 64 years (IQR 56.5-72.0), and the residents that they cared for were also mostly female (64.4%, n=65). Residents had been living in the nursing home for ≤ 1-year (28%, n=27), for 2-4 years (54%, n=52), and for ≥ 5 years (18%, n=17). Most residents had either Alzheimer’s dementia (52.5%, n=53), vascular dementia (18.8%, n=19), or another type or combination (29.4%, n=30).

### Activities in nursing homes

Participants reported a median of 20 distinct activities (IQR 12-24). The reported number of activities differed by participant group. Professionals reported a median of 33 activities (IQR 23-42), volunteers a median of 18 activities (IQR 14-27), and informal caregivers a median of 15 activities (IQR 7.5-20.5).

In total, 129 activities were mentioned (Table 2). The most frequently mentioned categories were music and entertainment (n=190), exercising (n=167), celebrating holidays and events (n=147), cognitive stimulation (n=141), and walking (n=141). Aromatherapy (n=32), sensory stimulation activities (n=51), activities including dolls (n=53), animal activities (n=87), and daily rituals (n=92) were the least mentioned categories.

**Table 2.** Overview of activities offered in nursing homes and their contribution to quality of life (n=207)

| Activities | Number of participants indicating the activity is offered (%) | Number of participants indicating the activity contributes to quality of life<sup>§</sup> |
| --- | --- | --- |
| **Music and entertainment** | 190 (91.8) |  |
| Live music and concerts | 144 (69.6) | 135 |
| Listening to music (cassette, cd, mp3, etc.) | 133 (64.3) | 128 |
| Watching TV/movies | 120 (58.0) | 91 |
| Singing | 119 (57.5) | 115 |
| Listening to the radio | 60 (29.0) | 51 |
| Playing a musical instrument | 30 (14.5) | 27 |
| Other | 1 (0.5) | 1 |
| **Exercise** | 167 (80.7) |  |
| (Duo-)cycling | 106 (51.2) | 97 |
| Dancing | 69 (33.3) | 65 |
| (Chair) yoga | 38 (18.4) | 33 |
| (Chair) gymnastics | 16 (7.7) | 14 |
| Other | 12 (5.8) | 12 |
| **Activities focused on holidays and events** | 147 (71.0) |  |
| Celebrating national holidays | 129 (62.3) | 113 |
| Celebrating birthdays | 122 (58.9) | 108 |
| Celebrating religious holidays | 103 (49.8) | 93 |
| Celebrations related to sporting events (e.g. ice skating, soccer, Olympics) | 81 (39.1) | 68 |
| Other | 3 (1.4) | 3 |
| **Cognitive stimulation** | 141 (68.1) |  |
| Reading a newspaper/ book/magazine | 88 (42.5) | 79 |
| Doing jigsaw/other puzzles | 86 (41.5) | 73 |
| Reading out loud | 75 (36.2) | 68 |
| Concentration games | 61 (29.5) | 49 |
| Card games | 37 (17.9) | 33 |
| Using the computer | 21 (10.1) | 20 |
| Writing | 10 (4.8) | 9 |
| Other | 5 (2.4) | 4 |
| **Walking (indoors and outside)** | 141 (68.1) | 137 |
| **Reminiscence** | 137 (66.2) |  |
| Retrieving and talking about memories | 125 (60.4) | 119 |
| Looking at photo albums | 97 (46.9) | 93 |
| Looking at photographs of children and babies | 54 (26.1) | 52 |
| Looking at/reading history books | 37 (17.9) | 35 |
| Other | 12 (5.8) | 10 |
| **Family and social activities** | 135 (65.2) |  |
| Drinking coffee together | 115 (55.6) | 111 |
| Eating together | 108 (52.2) | 100 |
| Family visits | 93 (44.9) | 89 |
| Play shuffleboard | 84 (40.6) | 68 |
| Watching an activity | 67 (32.4) | 58 □ |
| Telephoning someone | 63 (30.4) | 56 |
| Playing bingo | 59 (28.5) | 47 |
| Contact with (young) children | 44 (21.3) | 42 |
| Group conversations | 43 (20.8) | 39 |
| Shopping | 32 (15.5) | 29 |
| Helping others | 24 (11.6) | 24 |
| Other | 4 (1.9) | 3 |
| **Activities focused on personal care** | 131 (63.3) |  |
| Polishing nails/nail care | 101 (48.8) | 90 |
| Going to the hairdresser | 94 (45.4) | 81 |
| Combing hair | 90 (43.5) | 80 |
| Taking a bath/shower | 85 (41.1) | 77 |
| Putting on make-up | 41 (19.8) | 40 |
| Blow drying hair | 32 (15.5) | 30 |
| Other | 8 (3.9) | 8 |
| **Household activities** | 126 (60.9) |  |
| Setting table | 99 (47.8) | 91 |
| Folding laundry | 75 (36.2) | 65 |
| Cooking | 53 (25.6) | 49 |
| Baking | 52 (25.1) | 51 |
| Doing dishes | 51 (24.6) | 48 |
| Chores around the house | 11 (5.3) | 11 |
| Other | 7 (3.4) | 7 |
| **Outside activities** | 106 (51.2) |  |
| (Group) outings | 73 (35.3) | 69 |
| Take care of plants | 41 (19.8) | 37 |
| Gardening | 37 (17.9) | 30 |
| Bird watching | 34 (16.4) | 30 |
| Other | 9 (4.3) | 8 |
| **Art activities** | 101 (48.8) |  |
| Painting | 66 (31.9) | 54 |
| Drawing | 65 (31.4) | 49 |
| Arts and crafts | 64 (30.9) | 49 |
| Needle work (knitting, spool knitting, embroidering, crocheting) | 55 (26.6) | 39 |
| Looking at art | 21 (10.1) | 21 |
| Drama/theatre | 7 (3.4) | 7 |
| Poetry | 5 (2.4) | 5 |
| **Religious activities** | 101 (48.8) |  |
| Attending a religious meeting | 95 (45.9) | 74 |
| Praying | 41 (19.8) | 40 |
| Other | 1 (0.5) | 1 |
| **Activities focused on daily rituals** | 92 (44.4) |  |
| Getting the newspaper in the morning | 70 (33.8) | 67 |
| Drinking a glass of wine in the evening | 55 (26.6) | 52 |
| Other | 14 (6.8) | NASK |
| **Animal activities** | 87 (58.0) |  |
| Playing with and taking care of live animals | 51 (24.6) | 45 |
| Playing with and taking care of robotic animals | 42 (20.3) | 38 |
| Playing with and taking care of stuffed animals | 41 (19.8) | 34 |
| Other | 2 (1.0) | 2 |
| **Activities including dolls** | 53 (25.6) | 42 |
| **Sensory stimulation** | 51 (29.5) |  |
| Listening to nature sounds | 31 (15.0) | 26 |
| Tasting different flavours | 30 (14.5) | 28 |
| Massage | 29 (14.0) | 29 |
| Smelling different scents | 28 (13.5) | 24 |
| Snoezelen | 27 (13.0) | 25 |
| Sorting objects | 20 (9.7) | 16 |
| Use of activity cushion | 16 (7.7) | 13 |
| Feeling running water | 5 (2.4) | 3 |
| Other | 5 (2.4) | 2 |
| **Aromatherapy** | 32 (15.5) | 26 |
| Not knowing which activities are performed | 9 (4.3) | NA |

<sup>§</sup>This question is only answered by participants who reported the corresponding activity; <sup>||</sup>: $n = 206$ instead of 207, one participant did not complete all the questions; NASK: not asked in the survey; NA: not applicable.

When looking at specific activities, the top five most mentioned activities were live music and concerts (n=144, category music and entertainment), walking indoors or outside (n=141, category walking), listening to music (n=133, category music and entertainment), celebrating national holidays (n=129, category activities focused on holidays and events), and talking about memories (n=125, category reminiscence). The activities that were offered least frequently in nursing homes were chores around the house (n=11, category household), drama/theatre (n=7, category art), poetry (n=5, category art), writing (n=5), and feeling running water (n=5, category sensory stimulation).

### Expected contribution to quality of life

A median of 17 activities (IQR 9-31) were expected to contribute to the residents’ quality of life (see Table 2). Professionals expected a median of 30 activities (IQR 18-40) to contribute to the residents’ quality of life, volunteers a median of 16 (IQR 13-25), and informal caregivers a median of 10 (IQR 4-17). The median proportion of reported activities that were expected to contribute to the residents’ quality of life was 90% (IQR 75-97%). The professionals expected a higher proportion of activities to contribute to quality of life (Mdn=95.3, IQR 88.8-97.3) compared to informal caregivers (Mdn=80, IQR 66.7-94.5), and volunteers (Mdn=90, IQR 81.3-94.4).

Regarding the categories, Figure 2 shows that art activities, family and social activities, walking, music activities, and activities focused on personal care are most often expected to contribute to residents’ quality of life. By contrast, religious activities, activities including dolls, and aromatherapy were less often expected to contribute to residents’ quality of life. Still, at least 80% of the participants expected all categories to contribute to the residents’ quality of life.

**Figure 2.**
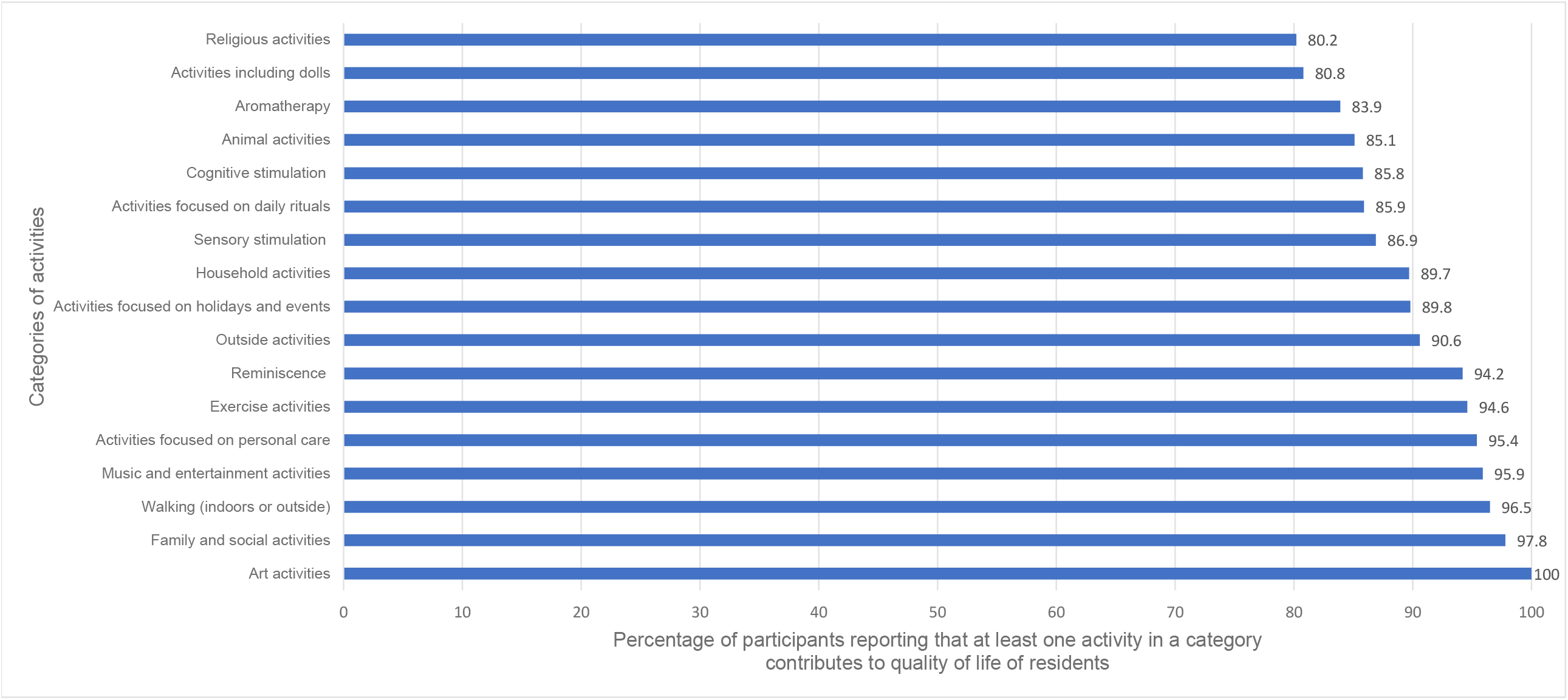
The expected contribution to residents’ quality of life per activity category

Looking at specific activities, massages (n=29), helping others (n=24), looking at art (n=21), chores around the house (n=11), drama/theatre (n=7), and poetry (n=5) were expected to contribute to the residents’ quality of life by all participants. By contrast, less than 80% of the participants expected some activities to contribute to quality of life, i.e., bingo (79.7%, n=59), attending religious gatherings (77.9%, n=95), arts and crafts (76.6%, n=64), watching movies/TV (75.8%, n=120), drawing (75.4%, n=65), needlework (70.9%, n=55), and feeling running water (60.0%, n=5). All other activities were expected to contribute to quality of life by at least 81% of the participants.

Finally, the mean score of participants on the extent to which activities were tailored to the individual’s wishes, needs, and skills was 6.5 (SD=1.9, n=193) on a VAS scale from 1(very bad) to 10 (very good). This score varied between participant groups, with a range of 6.4 for informal caregivers to 6.8 for volunteers.

### Choosing and performing activities

Healthcare assistants (134 times), informal caregivers (123 times), and occupational therapists (121 times) were most often involved in choosing and performing activities with residents (Figure 3). Managers, nurse practitioners, and physicians were least often reported as being involved in performing activities. Nurse practitioners, team leaders, and physicians were least mentioned for choosing activities.

**Figure 3.**
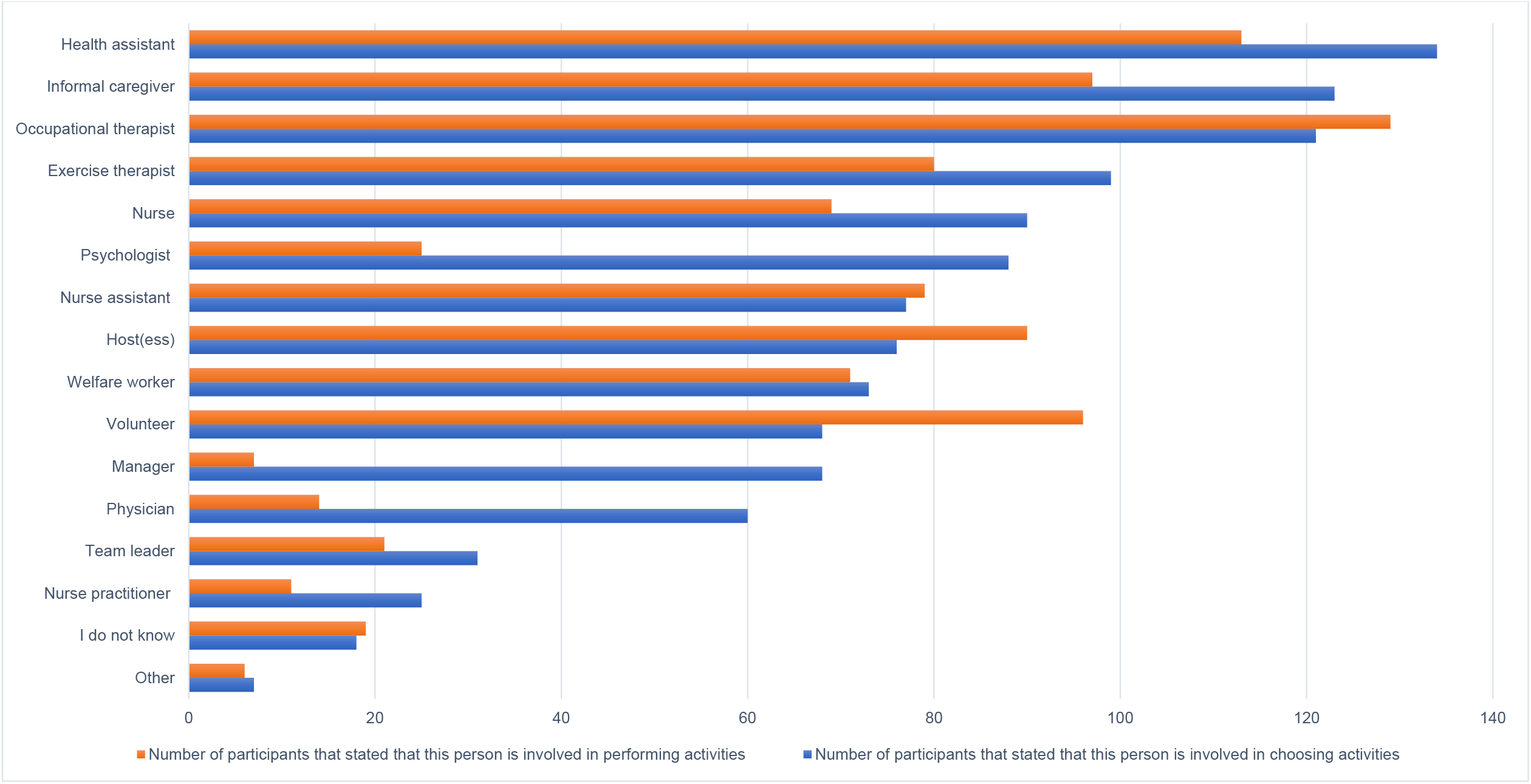
Participants involved in choosing or performing activities for nursing home residents with dementia.

## Discussion

This study shows that a wide range of activities are offered to persons living with dementia in nursing homes, and the most frequently offered activities relate to music and exercise. Most activities were expected by healthcare professionals, informal caregivers, and volunteers to contribute to residents’ quality of life. Occupational therapists, health assistants, and informal caregivers were most often involved in choosing and performing activities for nursing home residents with dementia.

This study found a discrepancy between the activity categories and specific activities within these categories. Because of the diversity of activities within a category it is not possible to make statements about activity categories (such as sensory stimulation activities are offered the least/most). This discrepancy is also visible in the expected contribution to residents’ quality of life in several categories, such as the music and entertainment category. For example, in accordance with previous research watching TV was mentioned frequently [21, 22]. However, fewer participants expected watching TV to contribute to the quality of life as compared to the overarching activity category music and entertainment. This finding illustrates the importance of selecting a specific activity, the expected contribution to quality of life varies within each activity category.

Studies looking at the effect of activities on quality of life of nursing home residents with dementia indicate that it matters which specific activity is offered to the resident to increase their quality of life [12, 14, 17, 23]. In our study, massages, helping others, looking at art, chores around the house, drama/theatre, and poetry were expected to contribute most to the residents’ quality of life. Interestingly, this expected contribution to quality of life is not always based on evidence; for instance, according to a mixed-methods longitudinal study with 125 residents no evidence for the impact of looking at art on quality of life was found in persons with dementia.[24] According to a between-groups quasi-experimental pretest–post-test design with 178 persons with dementia, performing drama also had no statistically significant effect on residents’ quality of life [25]. The effect of massages [26, 27] and watching TV on quality of life of nursing home residents needs further study. Hence, there is a gap between activities that are expected to contribute to the residents’ quality of life and activities that are actually shown to improve the residents’ quality of life in effect studies. This implies that the current organization of activities in Dutch nursing homes is probably more practice based than evidence based.

Interestingly, almost all offered activities were expected to contribute to residents’ quality of life, while the extent to which activities are being tailored to the individual resident was indicated to be rather low. This implies that quality of life may be improved by further tailoring activities to residents’ individual needs, interests, and abilities. This is also in line with aspects of the person-centered care standard for long-term care [28]. However, tailoring activities to residents’ needs can be challenging [29]. This could explain the relatively low score for tailoring activities to individual residents. Another explanation could be that it is easier to organize group activities that everyone can attend rather than tailored activities. To be able to tailor activities it is also important to know the person and to take time to discover their needs and preferences [5]. This can be done with the help of informal caregivers as they have knowledge about the resident’s (previous) preferences [30].

Our results show that informal caregivers are involved in choosing and performing activities, which is in line with earlier research that shows that informal caregivers contribute to many activities in nursing homes [31, 32]. Performing those activities together strengthens the relationship between a person with dementia and their loved ones [8]. In addition, the collaboration between informal caregivers, healthcare professionals, and volunteers may help the process of tailoring activities to the resident’s needs and preferences by combining their knowledge about the resident. This can be helpful, because this study shows that healthcare professionals and informal caregivers do not fully agree on which activities are expected to contribute to the residents’ quality of life.

This is the first nationwide study to summarize activities that are offered in nursing homes. The study provides insight into current activities and their expected contribution to residents’ quality of life. This overview of activities may inspire people to perform more tailored activities with nursing home residents with dementia, which is important as choosing activities can be difficult. Another strength of this study is the inclusion of perspectives of healthcare professionals as well as volunteers and informal caregivers.

A limitation of the study is that, due to the web-based nature of the survey, it is not possible to assess the non-response rate and the representativeness of our sample. Moreover, for some of the activities, the number of participants reporting on the expected contribution to quality of life was very small and therefore the observed contribution to quality of life of these activities may differ from the actual contribution in the overall population. Additionally, proxy reports were used instead of reports of residents themselves. This was the only practical option since the severity of the dementia of nursing home residents does not allow them to participate in a survey [33]. This may have affected the response, as family and staff are known to rate residents’ quality of life lower than residents themselves would do [33, 34]. Moreover, the perception of the definition of quality of life may differ between participants. The data were collected during the COVID-19 pandemic. This may have influenced the study, although previous research on activities during the pandemic showed that continuing individual activities was possible when adapted to the applicable guidelines and conditions, such as the use of other locations [35].

## 7. Conclusions

This study shows that many different activities are currently being offered to nursing home residents with dementia and not all activities are expected to contribute equally to the residents’ quality of life. Moreover, there is a discrepancy between activities expected to contribute to the quality of life and those offered most. The findings show that there is a gap between the contribution of activities to quality of life expected by caregivers and the evidence-based contribution of activities to quality of life. Activities should be tailored to both the residents’ needs and the available evidence about contribution to quality of life. Further research into the facilitators and barriers to offering tailored activities in collaboration with caregivers is necessary to unravel why and how tailoring activities should and can be improved.

## Data Availability

Due to the sensitive nature of the questions asked in this study, survey respondents were assured raw data would remain confidential and would not be shared.

## Acknowledgments

This work was supported by The Netherlands Organisation for Health Research and Development (ZonMw, grant 639003901). Funders had no role in the survey’s design, implementation, or analysis.

