## Supplementary Table S1 for "Activities contributing to quality of life of nursing home residents with dementia: a survey study"

**Supplementary Table S1.** Overview of the survey topics, questions and response options used

| Topic | Question number * | Question** | Number of items | Response options | Survey¶ |
|---|---|---|---|---|---|
| **1. Informed consent** | | | | | |
| | 1 | I want to participate in this study | 2 | Yes, no | H, V, I |
| | 2 | Which of the options below applies to you? | 2 | I work in a nursing home for residents with dementia, I have a loved one with dementia that lives in a nursing home, I volunteer in a nursing home for residents with dementia | H, V, I |
| **2. Demographics of the participant** [1] | | | | | |
| | 3 | What is your age? | 1 | Any number above 18 | H, V, I |
| | 4 | What is your gender? | 3 | Male, female, other | H, V, I |
| | 5 | Which province do you live in? | 13 | Noord-Holland, Zuid-Holland, Noord-Brabant, Zeeland, Overijssel, Utrecht, Limburg, Flevoland, Drenthe, Friesland, Groningen, Gelderland, I do not want to answer this question | H, V, I |
| | 6 | What is your country of birth? | 3 | The Netherlands, other, I do not want to answer this question | H, V, I |
| | 6.1 | Other | 197 | *All countries in the world* | H, V, I |
| | 7 | What is your father's country of birth? | 3 | The Netherlands, other, I do not want to answer this question | H, V, I |
| | 7.1 | Other | 197 | *All countries in the world* | H, V, I |
| | 8 | What is your mother's country of birth? | 3 | The Netherlands, other, I do not want to answer this question | H, V, I |
| | 8.1 | Other | 197 | *All countries in the world* | H, V, I |
| | 9 | What is your ideological background? | 9 | Protestant, Catholic, Islamic, Humanistic, Jewish, none, I do not want to answer this question, I do not know, other | H, V, I |
| | 9.1 | Other | 1 | Open question | H, V, I |
| | 10 | What is the highest level of education you have completed? | 9 | Primary education: < 6 years primary school, 6 years primary school, special primary education: lom-school (learning and behavioral difficulties), mlk-school (learning difficulties), More than primary school / primary school – no further completed education, vocational school, Secondary education: Mulo (more comprehensive lower education) / mms (secondary girls' school) / Mavo (intermediate general secondary education)/ vmbo (pre-vocational secondary education) / Mbo (secondary vocational education), Hbs /havo (senior general secondary education) / atheneum / gymnasium (pre-university education), HBO (higher professional education) / university, other, I do not know | H, V, I |
| | 10.1 | Other | 1 | Open question | H, V, I |

| | | | | | |
|---|---|---|---|---|---|
| 11 | What is your profession in the nursing home? | | 13 | Physician, psychologist, exercise therapist, health assistant, nurse assistent, nurse, nurse practitioner, welfare worker, host(ess), occupational therapist, team leader, manager, other | H |
| 11.1 | | Other | 1 | Open question | H |
| 12 | Do you have paid work? | | 2 | Yes, no | I |
| 12.1 | Yes … hours (if yes to paid work) | | 1 | Open question | I |
| 12.1.1 | Divided over … days (if yes to paid work) | | 1 | Open question | I |
| 12.2 | No … select which applies to you: (if no to paid work) | | 5 | Retired, no job because of illness or impairment, no job because of informal caregiving responsibilities, I am studying, other | I |
| 12.2.1 | | Other | 1 | Open question | I |
| 13 | Do you volunteer? | | 2 | Yes, no | I |
| 13.1 | Yes … hours (if yes to volunteer work) | | 1 | Open question | I |
| 13.1.1 | Divided over … days (if yes to volunteer work) | | 1 | Open question | I |
| **3. Demographics of the relative with dementia [1]** | | | | | |
| 14 | What is the age of your loved one? | | 1 | Any number above 18 | I |
| 15 | What is the gender of your loved one? | | 3 | Male, female, other | I |
| 16 | Which province does your loved one live in? | | 13 | Noord-Holland, Zuid-Holland, Noord-Brabant, Zeeland, Overijssel, Utrecht, Limburg, Flevoland, Drenthe, Friesland, Groningen, Gelderland, I do not want to answer this question | I |
| 17 | In which country was your loved one born? | | 3 | The Netherlands, other, I do not want to answer this question | I |
| 17.1 | | Other | 197 | *All countries in the world* | I |
| 18 | In which country was the father of your loved one born? | | 3 | The Netherlands, other, I do not want to answer this question | I |
| 18.1 | | Other | 197 | *All countries in the world* | I |
| 19 | In which country was the mother of your loved one born? | | 3 | The Netherlands, other, I do not want to answer this question | I |
| 19.1 | | Other | 197 | *All countries in the world* | I |
| 20 | What is the ideological background of your loved one? | | 9 | Protestant, Catholic, Islamic, Humanistic, Jewish, none, I do not want to answer this question, I do not know, other | I |
| 20.1 | | Other | 1 | Open question | I |
| 21 | What is the highest level of education your loved one completed? | | 9 | Primary education: < 6 years primary school, 6 years primary school, special primary education: lom-school (learning and behavioral difficulties), mlk-school (learning difficulties), More than primary school / primary school – no further completed education, vocational school, Secondary education: Mulo (more comprehensive lower education) / mms (secondary girls' school) / Mavo (intermediate general secondary | I |

| | | | | | |
|---|---|---|---|---|---|
| | | | | education)/ vmbo (pre-vocational secondary education) / Mbo (secondary vocational education), Hbs /havo (senior general secondary education) / atheneum / gymnasium (pre-university education), HBO (higher professional education) / university, other, I do not know | |
| 21.1 | | Other | 1 | Open question | I |
| 22 | When (approximately) was your loved one diagnosed with dementia? | | 1 | Open question date XX-XX-XXXX | I |
| 23 | What type(s) of dementia does your loved one have? | | 7 | Alzheimer's, Vascular dementia, Lewy Body Dementia (incl. Parkinson dementia), frontotemporal dementia, combination Alzheimer's and vascular dementia, other, I do not know | I |
| 23.1 | | Other | | Open question | I |
| 24 | Since when does your loved one with dementia live in the nursing home? | | 1 | Open question date XX-XX-XXXX | I |
| **4. Relationship between participant and their relative with dementia** | | | | | |
| 25 | What is your relationship with your loved one with dementia? | | 7 | Spouse/partner, son/daughter (in law), grandchild, brother/sister, nephew/niece/cousin, mentor, other | I |
| 25.1 | | Other | 1 | Open question | I |
| 26 | Did you live with your loved one before they moved to the nursing home? | | 1 | Yes, no | I |
| 27 | On average, how many hours per week do you spend with your loved one? | | 1 | Open question | I |
| 28 | Are you the person who is responsible for most of the care for your loved one? | | 1 | Yes, no | I |
| 29 | How long have you been caring for your loved one? | | 1 | Open question date XX-XX-XXXX | I |
| 30 | How many hours per month (on average) do you volunteer in a nursing home with residents with dementia? | | 1 | Open question | V |
| **5. Activities[2-4]** | | | | | |
| 31 | What do you think are the top five most common activities offered to people with dementia in the nursing home? | | 5 | Open question | H, V, I |
| 32 | What activities are offered to residents with dementia in the nursing home where you work? | | 18 | Multiple responses: Art activities, animal activities, activities including dolls, exercise, sensory stimulation, music and entertainment, reminiscence, outside activities, family and social activities, household activities, religious activities, activities focused on personal care, activities focused on holidays and events, aromatherapy, activities focused on daily rituals, others, I do not know | H, V, I |

| | | | | | |
|---|---|---|---|---|---|
| 32.1 | | Other | 1 | Open question | H, V, I |
| 32.1.1 | In your opinion, does this activity you mentioned under ‘’other’’ contribute to the quality of life of residents with dementia? | | 1 | Yes, no, I do not know | H, V, I |
| 32.2 | In your opinion, does aromatherapy contribute to the quality of life of residents with dementia? | | 1 | Yes, no, I do not know | H, V, I |
| 32.3 | Which specific art activity(ies) is/are involved? | | 10 | Multiple response: painting, drawing, arts and crafts, needle work, looking at art, drama/theatre, poetry, photography, filming, other | H, V, I |
| 32.3.1 | | Other | 1 | Open question | H, V, I |
| 32.3.2 | In your opinion, does [the selected art activity] contribute to the quality of life of residents with dementia? | | Depends on activities selected in the previous question | Yes, no, I do not know | H, V, I |
| 32.4 | Which specific animal activity(ies) is/are involved? | | 4 | Multiple response: playing with and taking care of live animals, playing with and taking care of robot animals, playing with or taking care of stuffed animals, other | H, V, I |
| 32.4.1 | | Other | 1 | Open question | H, V, I |
| 32.4.2 | In your opinion, does [the selected animal activity] contribute to the quality of life of residents with dementia? | | Depends on activities selected in the previous question | Yes, no, I do not know | H, V, I |
| 32.5 | Which specific exercise activity(ies) is/are involved? | | 4 | Multiple response: (Duo-)cycling, dancing, (chair)yoga, other | H, V, I |
| 32.5.1 | | Other | 1 | Open question | H, V, I |
| 32.5.2 | In your opinion, does [the selected exercise activity] contribute to the quality of life of residents with dementia? | | Depends on activities selected in the previous question | Yes, no, I do not know | H, V, I |
| 32.6 | Which specific sensory stimulation activity(ies) is/are involved? | | 9 | Multiple response: listening to nature sounds, tasting different flavors, massage, smelling different scents, Snoezelen, sorting objects, use of activity cushion, feeling running water, other | H, V, I |

| | | | | | |
|---|---|---|---|---|---|
| 32.6.1 | | Other | 1 | Open question | H, V, I |
| 32.6.2 | In your opinion, does [the selected sensory stimulation activity] contribute to the quality of life of residents with dementia? | | Depends on activities selected in the previous question | Yes, no, I do not know | H, V, I |
| 32.7 | Which specific music and entertainment activity(ies) is/are involved? | | 7 | Multiple response: live music and concerts, listening to music, watching TV/movies, singing, listening to the radio, playing instruments, other | H, V, I |
| 32.7.1 | | Other | 1 | Open question | H, V, I |
| 32.7.2 | In your opinion, does [the selected music and entertainment activity] contribute to the quality of life of residents with dementia? | | Depends on activities selected in the previous question | Yes, no, I do not know | H, V, I |
| 32.8 | Which specific cognitive stimulation activity(ies) is/are involved? | | 8 | Multiple response: reading a newspaper/book/magazine, doing jigsaws/puzzle, reading out loud, concentration games, card games, using the computer, writing, other | H, V, I |
| 32.8.1 | | Other | 1 | Open question | H, V, I |
| 32.8.2 | In your opinion, does [the selected cognitive stimulation activity] contribute to the quality of life of residents with dementia? | | Depends on activities selected in the previous question | Yes, no, I do not know | H, V, I |
| 32.9 | Which specific reminiscence activity(ies) is/are involved? | | 6 | Multiple response: talking about memories, looking at photo albums, looking at photographs of children and babies, looking at/reading history books, other | H, V, I |
| 32.9.1 | | Other | 1 | Open question | H, V, I |
| 32.9.2 | In your opinion, does [the selected reminiscence activity] contribute to the quality of life of residents with dementia? | | Depends on activities selected in the previous question | Yes, no, I do not know | H, V, I |
| 32.10 | Which specific outside activity(ies) is/are involved? | | 5 | Multiple response: (group) outing, taking care of plants, gardening, watching birds, other | H, V, I |

| | | | | | |
|---|---|---|---|---|---|
| 32.10.1 | | Other | 1 | Open question | H, V, I |
| 32.10.2 | In your opinion, does [the selected outside activity] contribute to the quality of life of residents with dementia? | | Depends on activities selected in the previous question | Yes, no, I do not know | H, V, I |
| 32.11 | Which specific family and social activity(ies) is/are involved? | | 12 | Multiple response: drinking coffee together, eating together, family visits, play shuffleboard, watching an activity, telephoning someone, playing bingo, contact with (young) children, group conversations, shopping, helping others, other | H, V, I |
| 32.11.1 | | Other | 1 | Open question | H, V, I |
| 32.11.2 | In your opinion, does [the selected family and social activity] contribute to the quality of life of residents with dementia? | | Depends on activities selected in the previous question | Yes, no, I do not know | H, V, I |
| 32.12 | Which specific household activity(ies) is/are involved? | | 7 | Multiple response: setting table, folding laundry, cooking, baking, doing dishes, chores around the house, other | H, V, I |
| 32.12.1 | | Other | 1 | Open question | H, V, I |
| 32.12.2 | In your opinion, does [the selected household activity] contribute to the quality of life of residents with dementia? | | Depends on activities selected in the previous question | Yes, no, I do not know | H, V, I |
| 32.13 | Which specific religious activity(ies) is/are involved? | | 3 | Multiple response: visiting a religious gathering, praying, other | H, V, I |
| 32.13.1 | | Other | 1 | Open question | H, V, I |
| 32.13.2 | In your opinion, does [the selected religious activity] contribute to the quality of life of residents with dementia? | | Depends on activities selected in the previous question | Yes, no, I do not know | H, V, I |

| | | | | | |
|---|---|---|---|---|---|
| 32.14 | Which specific activity(ies) focused on personal care is/are involved? | | 7 | Multiple response: polishing nails/nail care, going to the hairdresser, brushing hair, taking a bath/shower, doing make-up, blow drying hair, other | H, V, I |
| 32.14.1 | | Other | 1 | Open question | H, V, I |
| 32.14.2 | In your opinion, does [the selected activity focused on personal care] contribute to the quality of life of residents with dementia? | | Depends on activities selected in the previous question | Yes, no, I do not know | H, V, I |
| 32.15 | Which specific activity(ies) focused on holidays and events is/are involved? | | 5 | Multiple response: celebrating national holidays, celebrating birthdays, celebrating religious gatherings, celebrations related to sports events, other | H, V, I |
| 32.15.1 | | Other | 1 | Open question | H, V, I |
| 32.15.2 | In your opinion, does [the selected activity focused on holidays and events] contribute to the quality of life of residents with dementia? | | Depends on activities selected in the previous question | Yes, no, I do not know | H, V, I |
| 32.16 | Which specific activity(ies) focused on daily rituals is/are involved? | | 3 | Multiple response: getting the newspaper in the morning, drinking a glass of wine in the evening, other | H, V, I |
| 32.16.1 | In your opinion, does [the selected activity] contribute to the quality of life of residents with dementia? | | Depends on activities selected in the previous question | Yes, no, I do not know | H, V, I |
| **6. Preferences of the participants' relative with dementia** | | | | | |
| 33 | With regard to activities, how are the wishes, needs and skills of residents with dementia identified? | | 8 | Multiple response: conversation, booklet, collage, games, physical examination, none, other, I do not know | H, V, I |
| 33.1 | | Other | 1 | Open question | H, V, I |
| 34 | How would you score the extent to which activities are tailored to the individiual wishes, needs and skills/capacities of residents within the organization that you work in? | | 1 | VAS-scale very bad – 10 very good | H, V, I |

**7. People involved in activities**

| | | | | |
|---|---|---|---|---|
| 35 | Who is involved in choosing activities for residents with dementia? | 16 | Multiple response: physician, psychologist, therapist, health assistant, nurse, nurse practitioner, informal caregiver, occupational therapist, nurse assistant, host(ess), welfare worker, volunteer, manager, team leader, I do not know, other | H, V, I |
| 36 | Who is involved in performing activities for residents with dementia? | 16 | Multiple response: physician, psychologist, therapist, health assistant, nurse, nurse practitioner, informal caregiver, occupational therapist, nurse assistant, host(ess), welfare worker, volunteer, manager, team leader, I do not know, other | H, V, I |

Note: *Questions and follow-up questions were numbered to clarify which follow-up questions the respondents were shown, depending on their answers. They do not correspond with the original survey. **The questions that are similar for all participant groups are formulated as for healthcare professionals.
¶ H = Healthcare professional, V = Volunteer, I = Informal caregiver.
